# Effect of a widespread reduction in treatment duration for group A streptococcal pharyngitis on outcomes and household transmission

**DOI:** 10.1101/2024.10.09.24315208

**Authors:** Max Bloomfield, Hamish Reed, Sue Todd, Koen van der Werff, Michelle Balm, Tim Blackmore

## Abstract

**Background:** the optimal treatment duration for group A streptococcal pharyngitis (GAS-P) is debated. Shorter courses appear inferior for pharyngeal GAS eradication, however the effect of this on household transmission is uncertain. In 2022 a laboratory reporting change drove reduced treatment durations for GAS-P in our region. This study sought to assess the effect of this on outcomes.

**Methods:** positive throat swab cultures (TSC) for GAS from two years pre-change until 21 months post-change were matched to antibiotic dispensing data. Logistic models were fitted to examine associations between treatment duration and 30-day repeat antibiotic treatment, repeat GAS-positive TSC, and hospitalisation with complications; 90-day incidence of rheumatic fever; 30-day incident household GAS-P cases.

**Results:** 865 patients pre-change and 1604 post-change were included. Pre-change 32.8% received ≤7 days treatment, versus 60.0% post-change (p<0.01). There were no significant differences across any outcome measure at a population level between periods. When the post-change period was examined specifically, no significant differences occurred for any outcome measure for patients receiving five- or seven-days of antibiotics versus ten-days. Patients receiving no antibiotics also had similar outcomes, except for significantly higher odds of 30-day household cases (aOR 2.93, 95%-CI 1.44-5.96, p<0.01).

**Conclusions:** shorter treatment durations driven by a change in laboratory reporting resulted in no detectable change in GAS-P outcomes, except for more common household transmission in those receiving no treatment.

## Introduction

Sore throat is a common presentation to general practice.^1^ It is often treated with antibiotics, despite most sore throats being viral in origin.^2^ This overprescribing has been an area of concern with regard to antimicrobial stewardship (AMS).^1^ When *Streptococcus pyogenes* (group A streptococcus, GAS) is isolated many guidelines recommend ten days of antibiotics, rather than shorter courses, however the optimal duration is debated.^3–5^ A number of systematic reviews have found shorter courses to be non-inferior for clinical cure, although the majority of included studies use more broad-spectrum agents such as cephalosporins or macrolides as short course therapy.^6–11^ However, a systematic review found that microbiological failure (i.e. failure to eradicate GAS from the throat) is more common with shorter courses,^12^ which is consistent with a more recent randomised trial that found similar treatment success with five versus ten days of oral penicillin but observed lower bacterial eradication with the shorter duration.^13^ GAS is well-known to spread in households and other close-contact settings,^14^ which is why importance has been placed on bacterial eradication, with the reasoning being persistence likely increases the risk of transmission. However, to our knowledge, the effect of shorter course therapy for GAS pharyngitis on household transmission has never been examined directly.

Unlike most other developed nations New Zealand (NZ) has a high incidence of acute rheumatic fever (ARF),^15^ which occurs almost exclusively in people of Māori or Pacific ethnicity who consequently are targeted for prompt diagnosis and treatment of GAS pharyngitis as part of a national programme.^16^ Conversely, in those without ethnicity risk-factors, ARF is rare and regional and national guidelines recommend minimising throat swabbing and antibiotic treatment.^17,18^ As an AMS initiative our laboratory altered the reporting of throat swab cultures (TSCs) isolating GAS in 2022 to encourage less antibiotic use and shorter treatment durations for those without risk factors for ARF. This had an immediate and widespread effect in our region, with the proportion of patients being dispensed ten days of antibiotics following GAS-positive TSC dropping from 63% to 37%, with shorter durations becoming more common.^19^ As a continuation of this quality improvement project, this before-after retrospective cohort study sought to assess the effect of this reduction in treatment durations on key outcome measures for GAS pharyngitis treatment in our population: treatment failure requiring further antibiotic therapy, microbiological failure, suppurative complications requiring hospitalisation, ARF, and household transmission of GAS.

## Materials and Methods

### Setting

Awanui Labs Wellington is the single provider of clinical microbiology testing for the greater Wellington region of NZ, serving a population of approximately 500,000. TSC in our laboratory is on non-selective 5% sheep blood agar, with beta-haemolytic colonies followed up. The target organisms are GAS and *Streptococcus dysgalactiae* subspecies *equisimilis*. Other organisms are rarely reported. Reporting of scanty growths is discouraged. All results require electronic sign-off by requesters, and results for community samples are not reported to requesters until all testing is finalised (i.e. microscopy, culture and susceptibilities, if required). The change to laboratory reporting of TSC came into effect on 21 September 2022.

### Analysis

Creation of the analysis dataset has been described previously;^19–21^ briefly, between 1 October 2020 and 30 June 2024 all community TSC results from our laboratory were matched to a dataset containing all community antibiotic dispensing events from our region, using the National Health Index number, which is a unique identifier given to all patients in NZ. Dispensing data were supplied from the National Pharmaceutical Collection, which records all community dispensing of prescribed antibiotics. This allowed the relationship between sample collection, result reporting and antibiotic dispensing to be determined at an individual patient level. The pre-change period was defined as sample collection before 21 September 2022 and the post change as after this. The following patients were included in the primary analysis: 1) all those who had a TSC reported as growing GAS; and 2) were not already on antibiotics at time of laboratory report (i.e. no dispensing or ongoing antibiotics from five days before TSC collection up until date of report). Repeat episodes in the same patient of TSC with GAS within 30 days of the index sample were excluded and not counted as separate cases in the primary analysis (but were still included in the outcomes analysis). Patients with risk factors for ARF (Māori or Pacific ethnicity and aged 3-35 years) were also excluded and were used as a comparator group because there was no change in treatment duration between time periods (the change in laboratory reporting of TSC did not target this group). The antibiotic treatment duration each patient received following TSC was determined by what was dispensed in the five days following TSC report. Antibiotics that are not used for treatment of pharyngitis, e.g. nitrofurantoin, were excluded from this determination (Supplementary Table S1 for full list).

The primary outcome measures were: 1) isolation of GAS from a further TSC between completion of the initial antibiotic course and day 30 post completion; 2) further antibiotic dispensing between completion of the initial antibiotic course and day 30 post completion, with the same non-pharyngitis antibiotics excluded; 3) any unplanned hospitalisation within 30 days of TSC collection; 4) hospitalisation with infection relating to the pharynx, head or neck within 30 days of TSC collection; 5) a notified diagnosis of ARF within 90 days of TSC collection; 6) isolation of GAS from TSC from another individual with the same street address between day five and 30 post treatment completion. For these measures, where a patient did not receive antibiotics for their index GAS positive TSC ‘completion of the initial antibiotic course’ was regarded as day of TSC report. The hospitalisation outcomes were for the only two acute admitting hospitals in our region and included visits to the Emergency Department without overnight admission.

Two analyses were performed: the first compared the primary outcome measures between the pre- and post-change time periods across the entire cohort; the second compared primary outcomes according to the specific treatment duration dispensed for those in the post-change period. The chi-squared test was used to compare categorical variables and the Mann Whitney U test for continuous variables. A logistic regression model was fitted for each analysis and outcome measure, with age, sex, ethnicity, NZ Deprivation Index 2013 (a measure of social deprivation assigned by domicile),^22^ collection of sample at urgent care centre and season sample was collected in added as covariates. Analysis was performed in Stata-17 (StataCorp, College Station, Texas). Hospital research committee approval was gained. Following screening Health and Disability Ethics Committees review was waived (2024 OOS 21470), as this project formed part of a quality-improvement audit cycle for the laboratory. The study was performed and reported according to the STROBE guidelines.^23^

## Results

Figure 1 shows the formation of the analysis cohort from all patients having TSC collected in each time period. There was a significant increase in TSC positivity for GAS between time periods for those not already dispensed antibiotics at time of report (5.5% vs 13.8%, p<0.01). A small proportion of patients in each time period had missing data for course duration, which was more common for patients dispensed a liquid-formulation antibiotic. Those with missing course duration did not differ statistically from those with complete data when stratified by antibiotic formulation (Supplementary Table S2), so data were deemed missing at random and were excluded from the analysis. Demographics of the pre- and post-change cohorts are shown in Table 1. The median age reduced from 20.1 to 18.3 years (p=0.02), with a greater reduction observed in the comparator group (17.2 to 12.1 years, p<0.01). The age distribution histograms are shown in Supplementary Figures S1a-b. In the post-change period there were a higher proportion of samples from patients living in areas of lesser social deprivation (median NZ Deprivation index 5 pre-change versus 4 post-change, p<0.01). The antibiotic agents used were similar across periods, although penicillin VK use increased and amoxicillin use decreased in the analysis cohort. Table 2 shows course durations and outcome measures across time periods. There was a reduction in median antibiotic course duration (10 days pre- versus 7 days post-change, p<0.01), whereas there was no change in the comparator cohort (median 10 days in both periods, p=0.70). Those receiving ten days of antibiotics reduced from 67.2% pre-change to 40.0% post-change (Figure 2), with shorter durations becoming more common, particularly five days (2.4% to 20.1% post-change). There were no significant differences across the six primary outcome measures in the multivariate logistic analyses comparing time periods (Table 2). There were no hospitalisations with infections relating to the pharynx, head or neck, or incident cases of ARF in either period in the analysis cohort. Table 3 shows the results of the multivariate logistic analyses comparing primary outcome measures according to the course durations received for patients in the post-change period. There were no significant differences in the adjusted odds ratios (aORs) for repeat GAS positive TSC within 30 days across the different treatment durations. There were no significant differences for repeat antibiotic courses within 30 days, other than for patients receiving no antibiotics initially who were more likely to receive further antibiotics (13.2% versus 11.4% in the ten days group, aOR 1.76, 95%-CI 1.20-2.58, p<0.01). Of note, in this group 15 of 62 (24.2%) who received antibiotics within 30 days did so on day six post TSC report, just outside the five day window for course length determination (Supplementary Figure S2b). There were very few unplanned hospitalisations within 30 days and no hospitalisations in any of the groups with infections relating to the pharynx, head or neck. There were no significant differences across treatment groups for incident household cases within 30 days, other than the no antibiotics group which had significantly higher incidence (5.5% versus 2.1% in the ten days group, aOR 2.93, 95%-CI 1.44-5.96, p<0.01). The antibiotic agents used in the different course duration groups were similar, other than the seven days group, in which amoxicillin was used more frequently (Supplementary Table S3).

**Table 1.** Demographic and clinical characteristics for patients in the analysis and comparator cohorts.

|  | Pre-change |  | Post-change |  | P |
| --- | --- | --- | --- | --- | --- |
|  | N. | % | N. | % |  |
| **Analysis cohort (patients without ARF risk factors<sup>a</sup>)** |  |  |  |  |  |
| Total number of samples | 865 |  | 1604 |  |  |
| Age (years), median (IQR) | 20.1 | **(14.9,25.5)** | 18.3 | **(8.4,35.4)** | 0.02 |
| Female | 548 | **63.4%** | 969 | **60.4%** | 0.08 |
| Ethnic group |  |  |  |  | <0.01 |
| European | 685 | **82.1%** | 1107 | **69.0%** |  |
| Māori | 15 | **1.8%** | 46 | **2.9%** |  |
| Pacific | 16 | **1.9%** | 33 | **2.1%** |  |
| Asian | 93 | **11.2%** | 241 | **15.0%** |  |
| Other/Unknown | 25 | **3.0%** | 177 | **11.0%** |  |
| NZ Deprivation Index 2013<sup>b</sup>, median (IQR) | 5 | **(2,8)** | 4 | **(1,7)** | <0.01 |
| Sample collected at urgent care centre<sup>c</sup> | 109 | **12.6%** | 198 | **12.3%** | 0.85 |
| Season sample collected in |  |  |  |  | 0.35 |
| Spring | 224 | **25.9%** | 470 | **29.3%** |  |
| Summer | 181 | **20.9%** | 319 | **19.9%** |  |
| Autumn | 236 | **27.3%** | 413 | **25.7%** |  |
| Winter | 224 | **25.9%** | 402 | **25.1%** |  |
| Commonest five antibiotics dispensed |  |  |  |  | 0.01 |
| Phenoxymethylpenicillin (Penicillin VK) | 417 | **64.4%** | 814 | **71.7%** |  |
| Amoxicillin | 165 | **25.5%** | 203 | **17.9%** |  |
| Erythromycin | 25 | **3.9%** | 33 | **2.9%** |  |
| Cefalexin | 20 | **3.1%** | 60 | **5.3%** |  |
| Roxithromycin | 9 | **1.4%** | 8 | **0.7%** |  |
| Other | 12 | **1.9%** | 17 | **1.5%** |  |
| **Comparator cohort (patients with ARF risk factors<sup>a</sup>)** |  |  |  |  |  |
| Total number of samples | 291 |  | 448 |  |  |
| Age (years), median (IQR) | 17.2 | **(10.7,22,4)** | 12.1 | **(8.2,22.0)** | <0.01 |
| Female | 173 | **59.5%** | 261 | **58.3%** | 0.69 |
| Ethnic group |  |  |  |  | 0.35 |
| Māori | 168 | **57.7%** | 274 | **61.2%** |  |
| Pacific | 123 | **42.3%** | 174 | **38.8%** |  |
| NZ Deprivation Index 2013<sup>b</sup>, median (IQR) | 7 | **(4,9)** | 7 | **(4,10)** | 0.12 |
| Sample collected at urgent care centre<sup>c</sup> | 46 | 15.8% | 72 | 16.1% | 0.92 |
| Season sample collected in |  |  |  |  | 0.01 |
| Spring | 55 | 18.9% | 121 | 27.0% |  |
| Summer | 56 | 19.2% | 96 | 21.4% |  |
| Autumn | 90 | 30.9% | 138 | 30.8% |  |
| Winter | 90 | 30.9% | 93 | 20.8% |  |
| Commonest five antibiotics dispensed |  |  |  |  | 0.06 |
| Phenoxymethylpenicillin (Penicillin VK) | 168 | 76.4% | 267 | 79.2% |  |
| Amoxicillin | 42 | 19.1% | 41 | 12.2% |  |
| Cefalexin | 2 | 0.9% | 12 | 3.6% |  |
| Erythromycin | 2 | 0.9% | 11 | 3.3% |  |
| Co-amoxiclav | 2 | 0.9% | 3 | 0.9% |  |
| Other | 4 | 1.8% | 3 | 0.9% |  |

<sup>a</sup> In New Zealand these are: Māori or Pacific ethnicity and aged 3-35 years.
<sup>b</sup> This is a measure of social deprivation assigned by domicile, with higher numbers indicating increasing levels of social deprivation.
<sup>c</sup> These are walk in centers for short notice general practice-level care where results are typically forwarded to the person's normal general practitioner
GAS, group A *Streptococcus* (*Streptococcus pyogenes*); TSC, throat swab culture

**Table 2.**
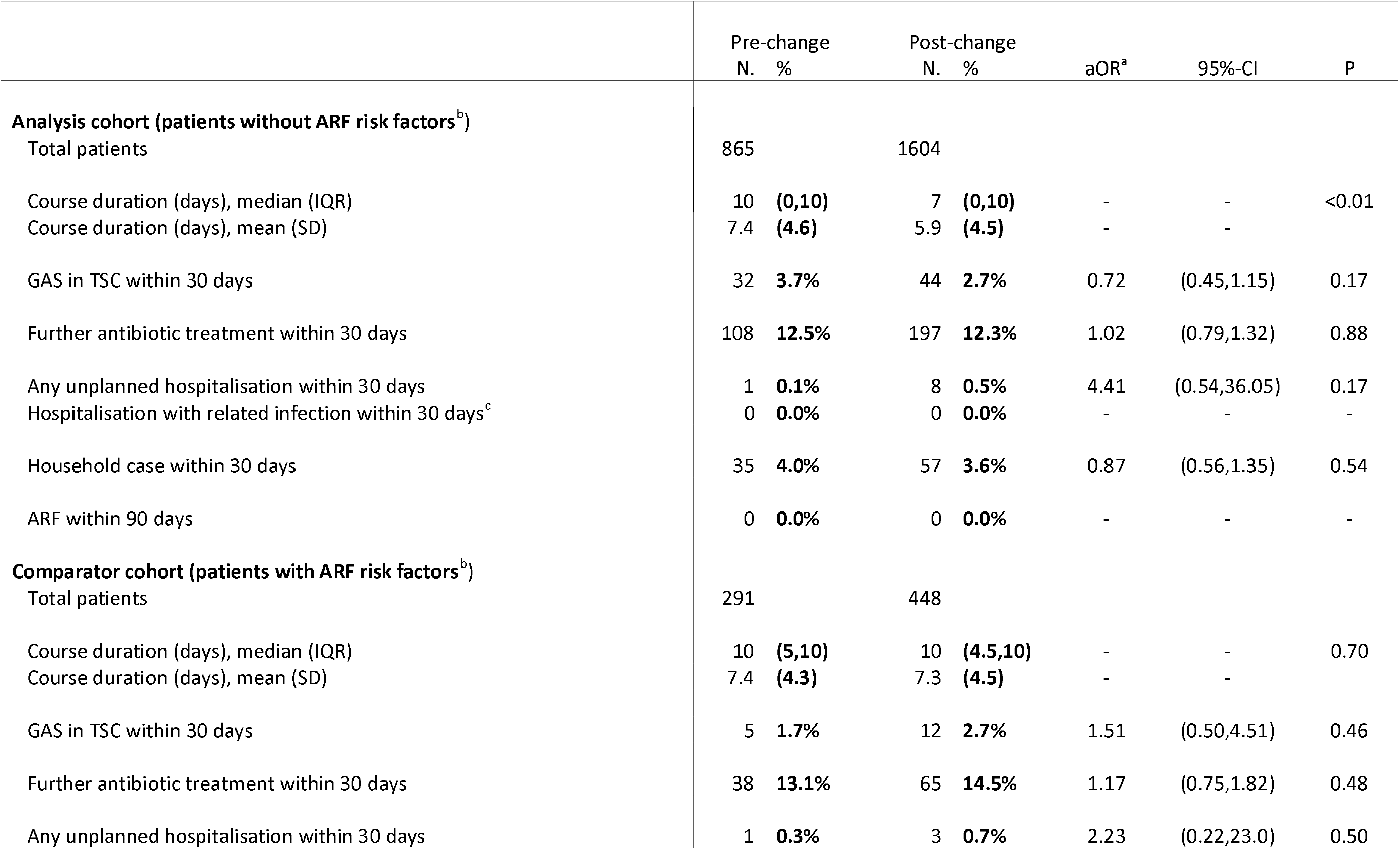
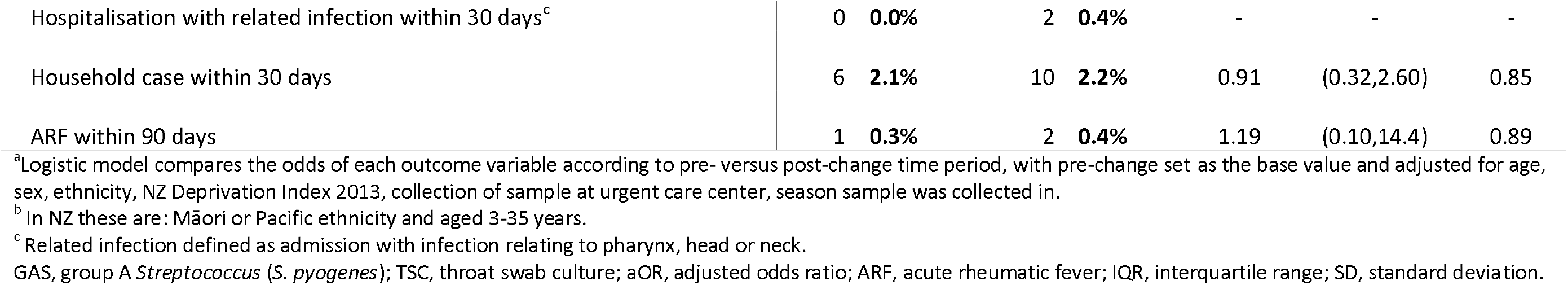
Antibiotic course durations and outcomes for patients in the analysis and comparator cohorts.

|  | Pre-change |  | Post-change |  | aOR<sup>a</sup> | 95%-CI | P |
| --- | --- | --- | --- | --- | --- | --- | --- |
|  | N. | % | N. | % |  |  |  |
| **Analysis cohort (patients without ARF risk factors<sup>b</sup>)** |  |  |  |  |  |  |  |
| Total patients | 865 |  | 1604 |  |  |  |  |
| Course duration (days), median (IQR) | 10 | (0,10) | 7 | (0,10) | - | - | <0.01 |
| Course duration (days), mean (SD) | 7.4 | (4.6) | 5.9 | (4.5) | - | - |  |
| GAS in TSC within 30 days | 32 | 3.7% | 44 | 2.7% | 0.72 | (0.45,1.15) | 0.17 |
| Further antibiotic treatment within 30 days | 108 | 12.5% | 197 | 12.3% | 1.02 | (0.79,1.32) | 0.88 |
| Any unplanned hospitalisation within 30 days | 1 | 0.1% | 8 | 0.5% | 4.41 | (0.54,36.05) | 0.17 |
| Hospitalisation with related infection within 30 days<sup>c</sup> | 0 | 0.0% | 0 | 0.0% | - | - | - |
| Household case within 30 days | 35 | 4.0% | 57 | 3.6% | 0.87 | (0.56,1.35) | 0.54 |
| ARF within 90 days | 0 | 0.0% | 0 | 0.0% | - | - | - |
| **Comparator cohort (patients with ARF risk factors<sup>b</sup>)** |  |  |  |  |  |  |  |
| Total patients | 291 |  | 448 |  |  |  |  |
| Course duration (days), median (IQR) | 10 | (5,10) | 10 | (4.5,10) | - | - | 0.70 |
| Course duration (days), mean (SD) | 7.4 | (4.3) | 7.3 | (4.5) | - | - |  |
| GAS in TSC within 30 days | 5 | 1.7% | 12 | 2.7% | 1.51 | (0.50,4.51) | 0.46 |
| Further antibiotic treatment within 30 days | 38 | 13.1% | 65 | 14.5% | 1.17 | (0.75,1.82) | 0.48 |
| Any unplanned hospitalisation within 30 days | 1 | 0.3% | 3 | 0.7% | 2.23 | (0.22,23.0) | 0.50 |
| Hospitalisation with related infection within 30 days<sup>c</sup> | 0 | 0.0% | 2 | 0.4% | - | - | - |
| Household case within 30 days | 6 | 2.1% | 10 | 2.2% | 0.91 | (0.32,2.60) | 0.85 |
| ARF within 90 days | 1 | 0.3% | 2 | 0.4% | 1.19 | (0.10,14.4) | 0.89 |

<sup>a</sup>Logistic model compares the odds of each outcome variable according to pre- versus post-change time period, with pre-change set as the base value and adjusted for age, sex, ethnicity, NZ Deprivation Index 2013, collection of sample at urgent care center, season sample was collected in.
<sup>b</sup> In NZ these are: Māori or Pacific ethnicity and aged 3-35 years.
<sup>c</sup> Related infection defined as admission with infection relating to pharynx, head or neck.
GAS, group A *Streptococcus* (*S. pyogenes*); TSC, throat swab culture; aOR, adjusted odds ratio; ARF, acute rheumatic fever; IQR, interquartile range; SD, standard deviation.

**Table 3.** Outcomes according to antibiotic course duration for patients in the post-change period analysis cohort.

|  | N. | % | aOR<sup>a</sup> | 95%-CI | P |
| --- | --- | --- | --- | --- | --- |
| **Total patients<sup>b</sup>** |  |  |  |  |  |
| Ten days | 612 | 40.0% | - | - | - |
| Seven days | 141 | 9.2% | - | - | - |
| Five days | 308 | 20.1% | - | - | - |
| No antibiotics | 470 | 30.7% | - | - | - |
| **GAS in TSC within 30 days** |  |  |  |  |  |
| Ten days | 22 | 3.6% | 1.00 | - | - |
| Seven days | 2 | 1.4% | 0.34 | (0.08,1.47) | 0.15 |
| Five days | 8 | 2.6% | 0.66 | (0.29,1.53) | 0.34 |
| No antibiotics | 9 | 1.9% | 0.48 | (0.20,1.14) | 0.10 |
| **Further antibiotic treatment within 30 days** |  |  |  |  |  |
| Ten days | 70 | 11.4% | 1.00 | - | - |
| Seven days | 16 | 11.3% | 0.96 | (0.54,1.74) | 0.92 |
| Five days | 39 | 12.7% | 1.13 | (0.74,1.72) | 0.59 |
| No antibiotics | 62 | 13.2% | 1.76 | (1.20,2.58) | <0.01 |
| **Any unplanned hospitalisation within 30 days** |  |  |  |  |  |
| Ten days | 5 | 0.8% | 1.00 | - | - |
| Seven days | 2 | 1.4% | 1.40 | (0.25,7.91) | 0.70 |
| Five days | 0 | 0.0% | - | - | - |
| No antibiotics | 0 | 0.0% | - | - | - |
| **Household case within 30 days** |  |  |  |  |  |
| Ten days | 13 | 2.1% | 1.00 | - | - |
| Seven days | 6 | 4.3% | 2.05 | (0.76,5.55) | 0.16 |
| Five days | 9 | 2.9% | 1.33 | (0.55,3.20) | 0.52 |
| No antibiotics | 26 | 5.5% | 2.93 | (1.44,5.96) | <0.01 |

Hospitalisation with infection relating to pharynx, head or neck and acute rheumatic fever have been omitted from this table because there were no cases.
<sup>a</sup> Logistic model compares the odds of each outcome variable according to course duration adjusted for age, sex, ethnicity, NZ Deprivation Index 2013, collection of sample at urgent care center, season sample was collected in, with ten days antibiotic duration set as the base value.
<sup>b</sup> Total (1531) is lower than 1604 from previous table due to a small number of patients being treated with course durations other than 0, 5, 7 or 10 days.
TSC, throat swab culture; GAS, group A *Streptococcus* (*Streptococcus pyogenes*); aOR, adjusted odds ratio; IQR, interquartile range; SD, standard deviation.

**Figure 1.**
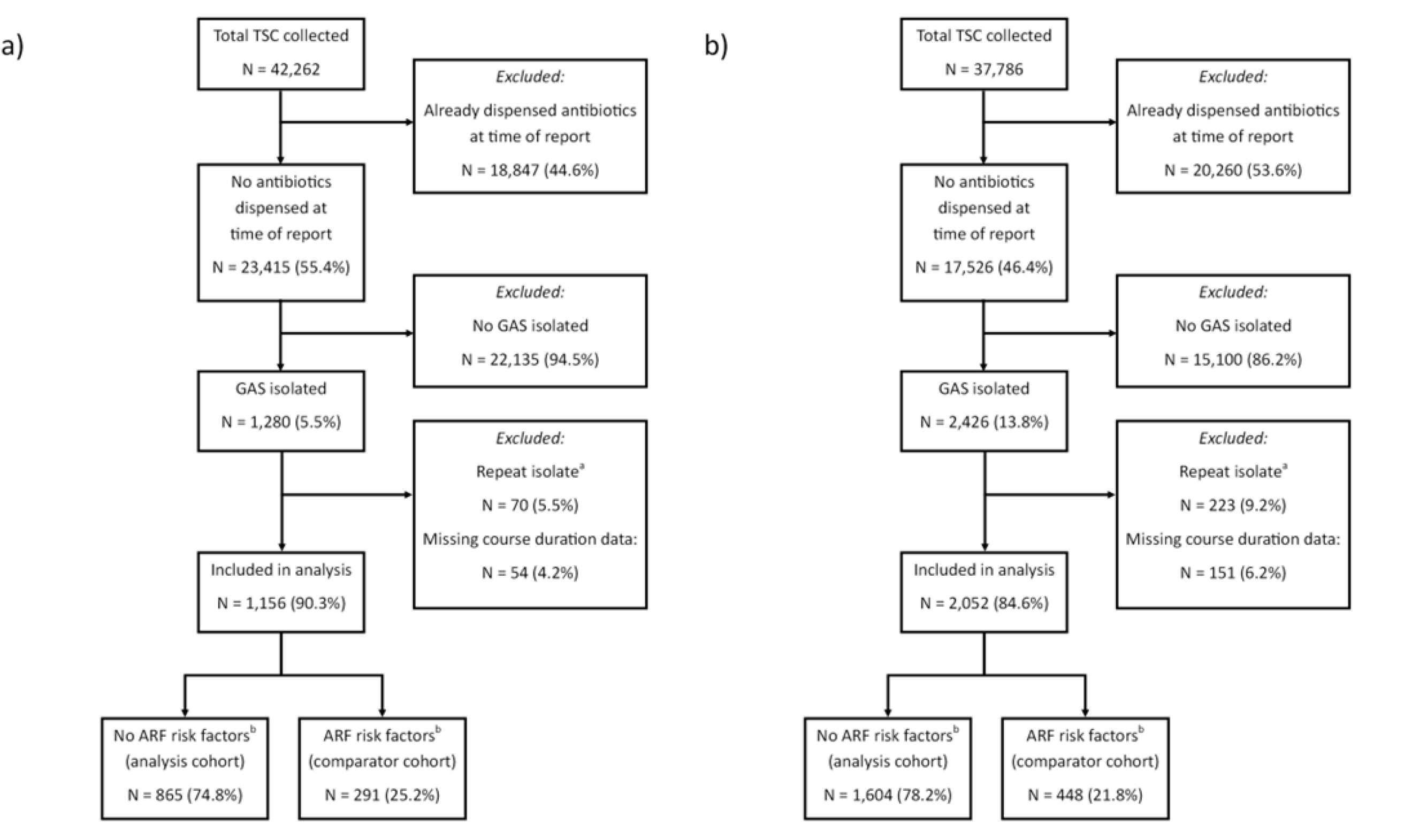
Formation of the analysis and comparator cohorts for a) pre-change period and b) post-change. ^a^Repeat positive throat swab cultures from the same patient within 30 days were excluded from the analysis. ^b^In New Zealand these are Māori or Pacific ethnicity and aged 3-35 years. TSC, throat swab culture; GAS, group *A Streptococcus*; ARF, acute rheumatic fever.

**Figure 2.**
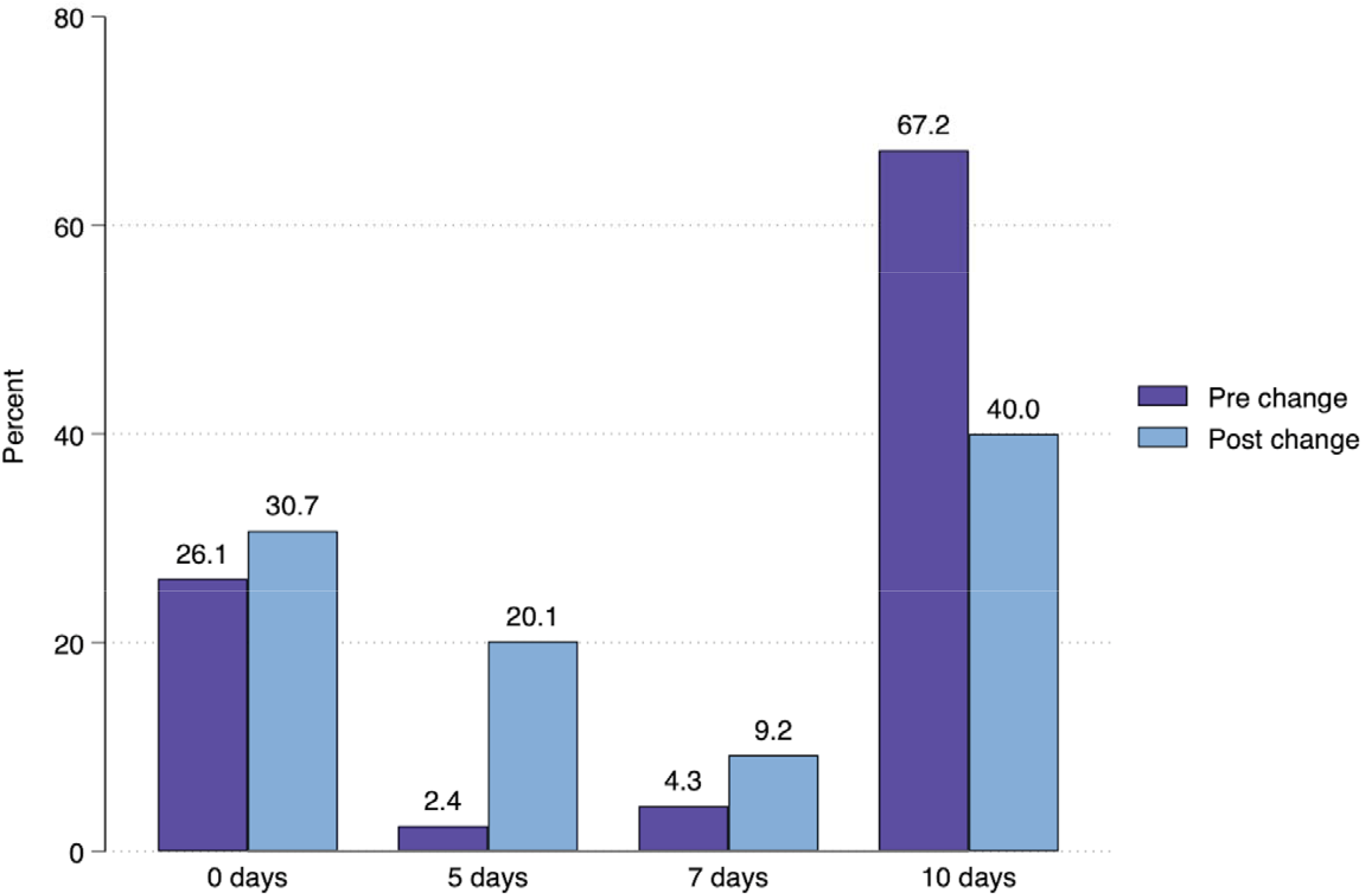
Percentage of patients receiving a given antibiotic course duration following positive throat swab culture for group *A Streptococcus* in the pre- and post-change time periods.

## Discussion

In this analysis key outcome variables for patients with GAS pharyngitis were examined across two time periods, between which there was an abrupt overall reduction in the antibiotic treatment durations used in our region. Because the change in course durations was driven by a change in laboratory reporting^19^ patients receiving shorter antibiotic courses in the post-change period represent individuals who would otherwise likely have been prescribed ten days of antibiotics.

Despite the shorter course durations, there was no signal from the data of increased risk of poor patient outcomes or household transmission, irrespective of duration of short-course antibiotics given. Those receiving no antibiotics had no increased incidence of further GAS positive TSC or hospitalisation but had an almost three-fold increase in the odds of a household case within 30 days. Although the no antibiotics group had significantly more 30-day antibiotic treatment, the difference appears to be driven largely by patients receiving antibiotics on day six post TSC report, which likely represents delayed dispensing from the original TSC report rather than treatment failure per se.

The findings from this study are consistent with prior evidence demonstrating similar treatment efficacy with shorter course antibiotic treatment of GAS pharyngitis. Much of this literature used agents other than penicillin or amoxicillin,^6–11^ so this study adds to recent trial evidence showing equivalent clinical outcomes using shorter-course penicillin-based treatment.^13^ Shorter course treatment has been associated with poorer pharyngeal GAS eradication, with most studies assessing this via programmed repeat TSC at a set time point after treatment.^12^ The clinical relevance of ongoing TSC positivity for GAS in patients that have clinically recovered is less certain, and to our knowledge this study is the first time the effect of shorter course therapy on real-world household transmission of GAS has been examined. Here we found no difference in incident household cases of GAS pharyngitis in those receiving shorter course treatment, but a significant increase in those receiving no antibiotics. This may indicate that short course treatment provides sufficient GAS suppression to limit household transmission, despite lower bacterial eradication when compared to ten days of antibiotics, whereas no antibiotic treatment does not.

Strengths of this study include a large sample size, important in the context of a study where most outcome measures were found not to differ between groups. Despite the large sample size, matching of laboratory and antibiotic treatment data occurred at the individual patient level, meaning these individual relationships could be precisely defined at scale rather than needing to be aggregated at a population level, as is common with observational studies. The nature of our local health services, with a single lab provider and only two acute admitting hospitals, also meant that we can be confident of near complete coverage and follow-up for outcome measures. We also examined multiple different potential indicators of treatment failure, increasing the sensitivity of the analysis for detecting possible suboptimal outcomes due to shorter treatment durations. Finally, a control group in which treatment durations did not change was available for comparison, which demonstrated that outcome measures were otherwise stable over time.

Limitations of this study include its observational nature and the potential for residual confounding after adjustment of the logistic models for measurable potential confounders. As such, differences in treatment duration at the individual patient level could have been influenced by unmeasured patient factors that could also influence outcome e.g. clinicians may have felt more comfortable using shorter treatment durations in patients with milder symptoms at baseline, who may also have been less likely to have treatment failure. Outcome measures were also determined via passively collected data, based on re-presentation to healthcare services, rather than systematic follow-up, as would occur in a trial setting. However, this may be both a limitation and a strength, as re-presentation is a demonstration of an outcome that was sufficiently important to the patient to seek further help. This may be a better reflection of the real-world effects of shorter course durations. For example, we did not collect TSC on household members of those being treated for GAS pharyngitis to determine the true incidence of household transmission. However, transmission leading to a laboratory diagnosis of GAS pharyngitis in a household member, as in this study, is likely a more relevant outcome. In this study we only examined patients who were not on antibiotics at the time of TSC result reporting, which represented 55.4% and 46.4% of all patients having TSC collected pre- and post-change, respectively (Figure 1). This was because there was a lesser reduction in treatment durations in those prescribed antibiotics pre-lab report across time periods (because the lab report itself is what drove the shorter durations).^19^ It is possible that patients included in this study may therefore represent patients with milder symptoms, as clinicians felt comfortable awaiting laboratory results before prescribing. Finally, this study was performed in a single geographical region, so results may not necessarily be generalisable to other populations, however the population in our region is relatively heterogeneous and likely similar to many populations in other developed countries.

In this cohort a widespread reduction in antibiotic treatment duration for GAS pharyngitis driven by changes in laboratory reporting does not appear to have resulted in detectable patient harm or increased household transmission of GAS. These outcomes were seen at a population level between time periods, and at an individual course duration level. Many patients did not receive any antibiotic treatment and did not appear to come to harm, suggesting that not all GAS pharyngitis necessarily requires antibiotic treatment with respect to individual patient risk, however there was a clear increase in household cases if no treatment was given initially. Although five- or seven-day treatment durations have been associated with lower bacterial eradication rates previously, we could find no evidence in this real-world setting that they result in an increase in subsequent household cases of GAS pharyngitis. This is a novel finding as far as we are aware.

## Data Availability

All data produced in the present work are contained in the manuscript.

## Acknowledgements

The authors would like to thank Te Whatu Ora – Ministry of Health for help in accessing the National Pharmaceutical Collection data, and Peter Wash and Joe Hart for help in accessing demographic and laboratory data.

## Transparency declaration

The authors declare that they have no conflicts of interest.

