## Supplementary Figures for "Effect of a widespread reduction in treatment duration for group A streptococcal pharyngitis on outcomes and household transmission"

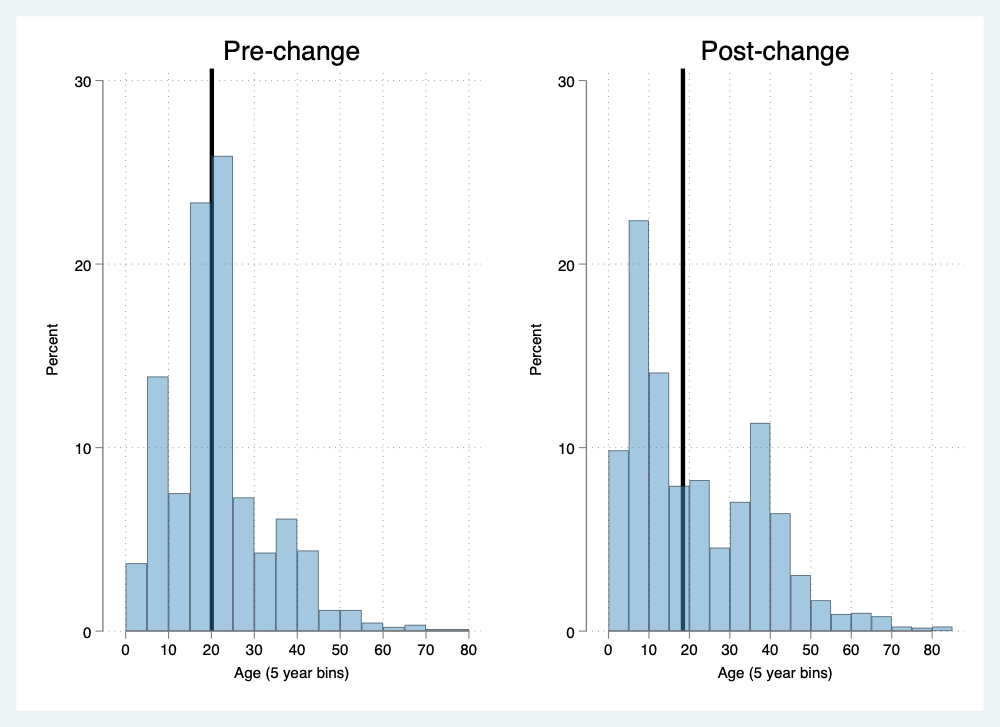

Figure S1a. Age distribution histograms for the analysis cohort (no risk factors for acute rheumatic fever)

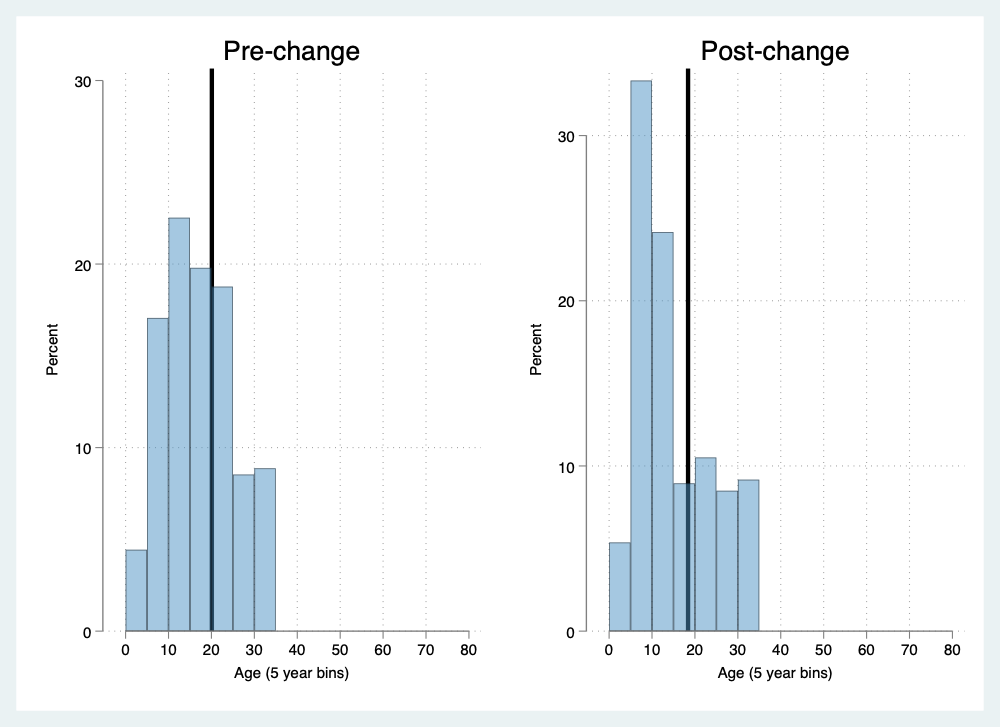

Figure S1b. Age distribution histograms for the comparator cohort (risk factors for acute rheumatic fever)

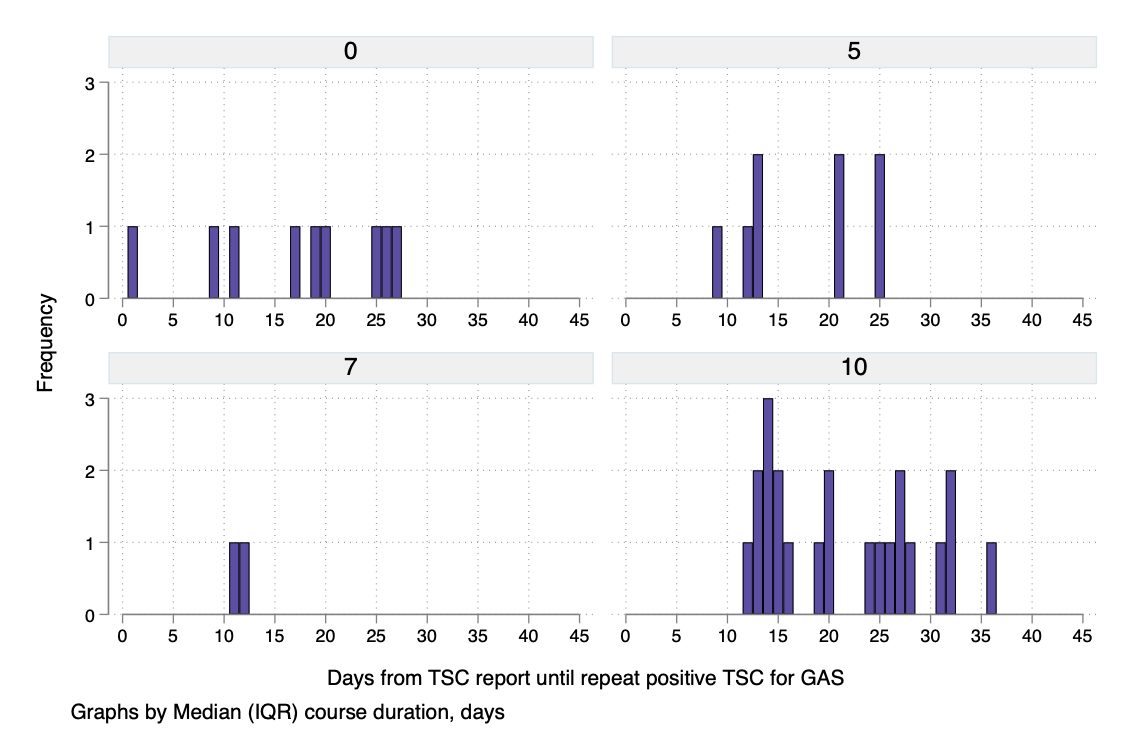

Figure S2a. Time from index throat swab culture report until repeat positive throat swab culture within 30 days according to initial treatment duration

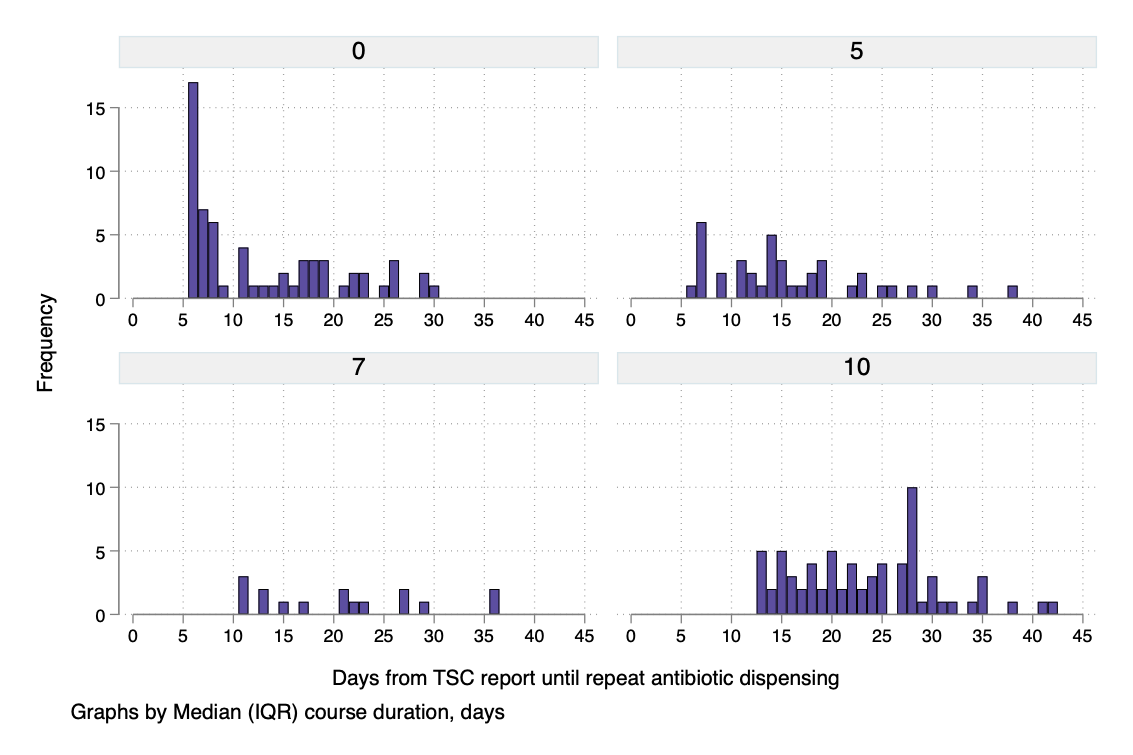

Figure S2b. Time from index throat swab culture report until repeat antibiotic dispensing within 30 days according to initial treatment duration

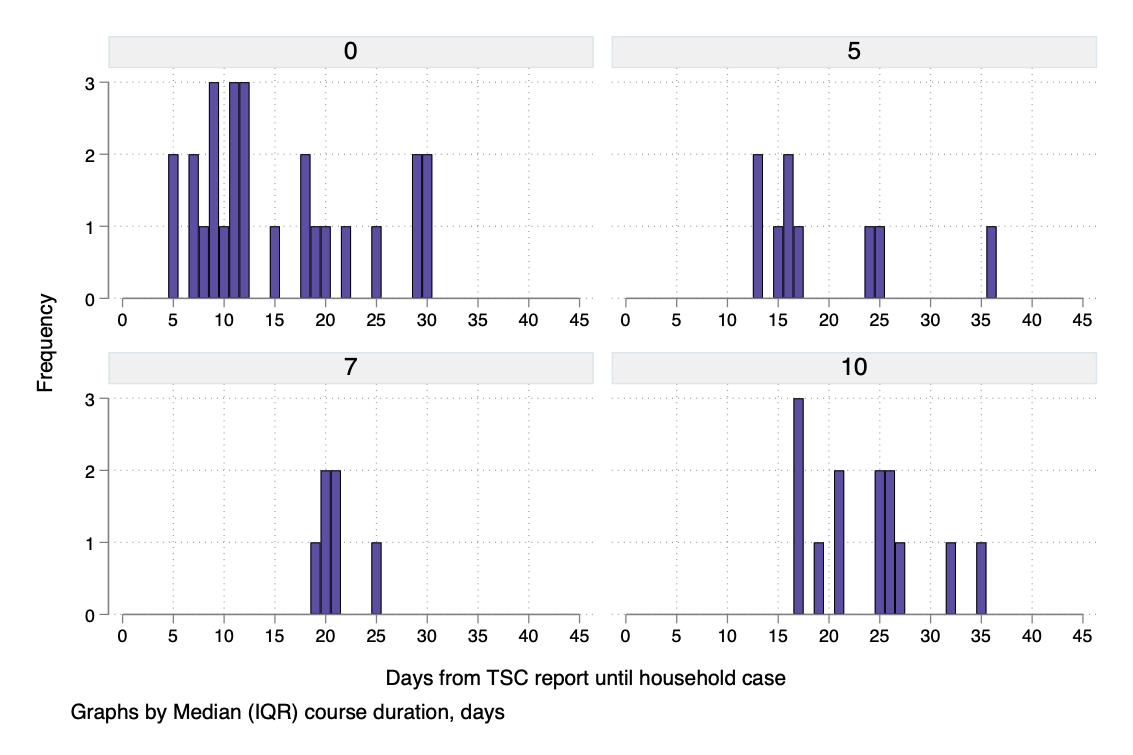

Figure S2c. Time from index throat swab culture report until household case within 30 days according to initial treatment duration
