## Supplementary Tables for "Effect of a widespread reduction in treatment duration for group A streptococcal pharyngitis on outcomes and household transmission"

Table S1. Non-pharyngitis antimicrobials excluded from the course duration and repeat antibiotics within 30 days determination

Agent

Ciprofloxacin

Metronidazole

Nitrofurantoin

Norfloxacin

Pivmecillinam

Trimethoprim

All topical antibiotics

All antifungals

| Supplementary Table S2. Demographic and clinical characteristics of patients with missing antibiotic course duration data | | | | | |
| --- | --- | --- | --- | --- | --- |
|  | Missing course duration | | | |  |
|  | No | | Yes | |  |
|  | N. | **%** | N. | % | P value |
| **Tablet/capsule formulation antibiotics** |  |  |  |  |  |
| Total number of samples | 1613 | **98.2%** | 29 | **1.8%** |  |
| Age (years), median (IQR) | 24.5 | **(19.0,37.4)** | 30.8 | **(22.2,40.4)** | 0.07 |
| Female | 1080 | **67.0%** | 19 | **65.5%** | 0.97 |
| Ethnic group |  |  |  |  | 0.86 |
| European | 1289 | **79.9%** | 25 | **86.2%** |  |
| Māori | 52 | **3.2%** | 0 | **0.0%** |  |
| Pacific | 43 | **2.7%** | 1 | **3.4%** |  |
| Asian | 181 | **11.2%** | 2 | **6.9%** |  |
| Other/Unknown | 48 | **3.0%** | 1 | **3.4%** |  |
| NZ Deprivation Index 2013^a^, median (IQR) | 5.0 | **(2,7)** | 5.0 | **(2,8)** | 0.48 |
| Sample collected at urgent care centre^b^ | 185 | **11.5%** | 4 | **13.8%** | 0.70 |
| Season sample collected in |  |  |  |  | 0.21 |
| Spring | 428 | **26.5%** | 5 | **17.2%** |  |
| Summer | 335 | **20.8%** | 6 | **20.7%** |  |
| Autumn | 450 | **27.9%** | 6 | **20.7%** |  |
| Winter | 400 | **24.8%** | 12 | **41.4%** |  |
| GAS in TSC within 30 days | 42 | **2.6%** | 1 | **3.4%** | 0.78 |
| Further antibiotic treatment within 30 days | 204 | **12.6%** | 4 | **13.8%** | 0.85 |
| Any unplanned hospitalisation within 30 days | 6 | **0.4%** | 0 | **0.0%** | 0.74 |
| Hospitalisation with related infection within 30 days | 0 | **0.0%** | 0 | **0.0%** | - |
| Household case within 30 days | 53 | **3.3%** | 0 | **0.0%** | 0.32 |
| ARF within 90 days | 0 | **0.0%** | 0 | **0.0%** | - |
| **Liquid formulation antibiotics** |  |  |  |  |  |
| Total number of samples | 704 | **85.4%** | 120 | **14.6%** |  |
| Age (years), median (IQR) | 7.4 | **(5.3,9.6)** | 6.9 | **(5.1,8.8)** | 0.12 |
| Female | 343 | **48.7%** | 53 | **44.2%** | 0.54 |
| Ethnic group |  |  |  |  | 0.23 |
| European | 503 | **71.4%** | 81 | **67.5%** |  |
| Māori | 9 | **1.3%** | 3 | **2.5%** |  |
| Pacific | 6 | **0.9%** | 0 | **0.0%** |  |
| Asian | 153 | **21.7%** | 28 | **23.3%** |  |
| Other/Unknown | 33 | **4.7%** | 8 | **6.7%** |  |
| NZ Deprivation Index 2013^a^, median (IQR) | 4.0 | **(1,7)** | 5.0 | **(1,6.5)** | 0.10 |
| Sample collected at urgent care centre^b^ | 105 | **14.9%** | 17 | **14.2%** | 0.83 |
| Season sample collected in |  |  |  |  | 0.38 |
| Spring | 196 | **27.8%** | 33 | **27.5%** |  |
| Summer | 138 | **19.6%** | 25 | **20.8%** |  |
| Autumn | 165 | **23.4%** | 35 | **29.2%** |  |
| Winter | 205 | **29.1%** | 27 | **22.5%** |  |
| GAS in TSC within 30 days | 32 | **4.5%** | 9 | **7.5%** | 0.17 |
| Further antibiotic treatment within 30 days | 101 | **14.3%** | 22 | **18.3%** | 0.26 |
| Any unplanned hospitalisation within 30 days | 3 | **0.4%** | 1 | **0.8%** | 0.74 |
| Hospitalisation with related infection within 30 days | 0 | **0.0%** | 0 | **0.0%** | - |
| Household case within 30 days | 32 | **4.5%** | 1 | **0.8%** | 0.06 |
| ARF within 90 days | 0 | **0.0%** | 0 | **0.0%** | - |
| Missing data have been divided up by antibiotic formulation type because liquid formulation antibiotics were more likely to have missing data.  ^a^ This is a measure of social deprivation assigned by domicile, with higher numbers indicating increasing levels of social deprivation.  ^b^ These are walk in centers for short notice general practice-level care where results are typically forwarded to the person’s normal general practitioner  IQR, interquartile range; GAS, group A *Streptococcus* (*Streptococcus* *pyogenes*); TSC, throat swab culture; ARF, acute rheumatic fever. | | | | | |

Table S3. Antibiotic agents used according to course duration for patients in the post-change period analysis cohort.

|  | N. | % |
| --- | --- | --- |
| Five-day course |  |  |
| Drug name |  |  |
| Phenoxymethylpenicillin (Penicillin VK) | 228 | **74.0%** |
| Amoxicillin | 48 | **15.6%** |
| Erythromycin | 3 | **1.0%** |
| Cefalexin | 23 | **7.5%** |
| Flucloxacillin | 1 | **0.3%** |
| Other | 5 | **1.6%** |
| Seven-day course |  |  |
| Drug name |  |  |
| Phenoxymethylpenicillin (Penicillin VK) | 78 | **55.7%** |
| Amoxicillin | 47 | **33.6%** |
| Erythromycin | 2 | **1.4%** |
| Cefalexin | 6 | **4.3%** |
| Flucloxacillin | 1 | **0.7%** |
| Other | 6 | **4.3%** |
| Ten-day course |  |  |
| Drug name |  |  |
| Phenoxymethylpenicillin (Penicillin VK) | 460 | **75.2%** |
| Amoxicillin | 87 | **14.2%** |
| Erythromycin | 27 | **4.4%** |
| Cefalexin | 28 | **4.6%** |
| Other | 10 | **1.6%** |

GAS, group A *Streptococcus* (*S. pyogenes*); TSC, throat swab culture.
